# MRI-Perivascular Spaces in Chronic and Episodic Migraine Disorder

**DOI:** 10.64898/2026.03.18.26348481

**Authors:** William Pham, Donggyu Rim, Alexander Jarema, Zhibin Chen, Mohamed Salah Khlif, Noemi Meylakh, Richard J. Stark, Amy Brodtmann, Vaughan G. Macefield, Luke A. Henderson

## Abstract

Migraine is a common and disabling neurological disorder linked to alterations in neuronal activity and waste clearance in the brain. MRI-visible perivascular spaces (PVS) are key components of the glymphatic system which may serve as imaging biomarker of such disorder. We hypothesised that higher frequency of migraine episodes would be associated with increased PVS burden, reflecting greater levels of impaired glymphatic clearance. In this retrospective case-control study of 90 participants (20 episodic migraineurs, 18 chronic migraineurs, and 52 age– and sex-matched healthy controls; 58 females, median [Q1, Q3] age=28.6 [25.1, 39.4] years) we investigated PVS alterations in episodic migraineurs (*n*=20) and 18 chronic migraineurs (*n*=18). PVS volumes and cluster counts were quantified in the white matter (WM), basal ganglia (BG), midbrain, and hippocampus. We stratified PVS metrics by white matter lobes and arterial vascular territories. After adjusting for age, sex, and total brain volume, episodic migraineurs exhibited significantly lower BG-PVS volumes (exp(β)=0.76, 95%CI [0.61, 0.94], *p*=0.01) compared to controls. Chronic migraineurs exhibited significantly lower PVS cluster counts in the parietal (exp(β)=0.8, 95%CI [0.68, 0.94], *p*=0.01) and temporal lobes (exp(β)=0.72, 95%CI [0.53, 0.96], *p*=0.03) and middle cerebral artery territory (exp(β)=0.82, 95%CI [0.68, 0.97], *p*=0.03) compared to healthy controls. Within migraineurs, those with aura (*n*=20) exhibited significantly lower PVS burden in all brain regions, vascular territories, and across the frontal, parietal, and temporal lobes (all *p*_FDR_<0.05). Our findings suggest that the aura symptom, rather than the migraine disorder itself, may primarily drive changes in perivascular spaces, with effects varying across brain regions.

## Introduction

Migraine is a highly prevalent and disabling neurological disorder, affecting over one billion individuals worldwide (Ferrari et al., 2022). It is characterised by recurring headache attack accompanied by a constellation of symptoms such as photophobia, phonophobia, allodynia, hyperalgesia, and nausea (Ferrari et al., 2022). The clinical presentation of migraine is highly heterogenous, with individuals experiencing distinct symptoms, frequencies, and severities. This complexity has contributed to the persistent challenge of fully elucidating the underlying pathophysiological mechanisms.

Recent evidence has implicated dysfunction of the glymphatic system, a brain-wide clearance pathway that facilitates the exchange of cerebrospinal fluid (CSF) and interstitial fluid (ISF), in migraine pathophysiology (Schain et al., 2017). A study of an animal model of migraine reported impaired glymphatic clearance combined with increased neural activity, reactive astrogliosis, and neuroinflammatory changes where trigeminal afferents first terminate in the spinal trigeminal nucleus (Huang et al., 2023). Furthermore, glymphatic inhibition exacerbated pain sensitivity during migraine-like episodes, suggesting a direct link between fluid clearance dysfunction and sensory processing abnormalities (Huang et al., 2023).

Approximately one-third of migraineurs experience aura, which is associated with cortical spreading depression (CSD) (Ayata & Lauritzen, 2015; Pietrobon & Moskowitz, 2014). Experimental evidence indicates that CSD induces a rapid collapse of perivascular spaces (PVS) at the cortical surface (Schain et al., 2017). Consequently, CSF influx along the glymphatic pathways is disrupted and downstream glymphatic transport and solute clearance in the parenchyma is stalled.

PVS are fluid-filled channels that line the cerebral vasculature serving and serve as the structural conduits for glymphatic flow. On MRI, they appear as CSF-isointense, tubular structures aligned with perforating vessels, and are typically less than 3 mm in diameter (Pham et al., 2022; Wardlaw et al., 2013). The enlargement of MRI-visible PVS is increasingly recognised as a potential biomarker of glymphatic dysfunction. A small number of MRI studies have reported associations between increased PVS burden and headaches or migraines. For example, enlarged PVS have been observed in children with headaches (Jeon et al., 2023) or migraines (Schick et al., 1999) compared to age-matched controls. Another study found greater PVS burden in the centrum semiovale and midbrain of adult migraineurs compared to controls (Yuan et al., 2023). However, a population study failed to observe significant associations between PVS enlargement in the white matter or basal ganglia with the presence of headaches and/or migraine (Husøy et al., 2016). Thus, the evidence regarding alterations in MRI-visible PVS associated with migraine and headache disorders is inconclusive.

Importantly, prior studies have relied on semi-quantitative visual rating scales for MRI-PVS measurements. To date, no studies have leveraged high-resolution 7T MRI and automated deep learning methods to precisely quantify PVS burden across multiple brain regions in MRI. Furthermore, the impact of migraine chronicity, episodic versus chronic, on PVS morphology remains unexplored. Given the increased allostatic burden, migraine frequency likely has a significant effect on glymphatic clearance and the presence of PVS on MRI. In this study, we aimed to characterise MRI-visible PVS alterations in individuals with episodic and chronic migraine using 7T MRI. We hypothesised that higher frequency of migraine episodes would be associated with increased PVS burden, reflecting greater levels of impaired glymphatic clearance.

## Methods

### Participants

A total of 90 participants were enrolled, comprising 20 episodic migraineurs, 18 chronic migraineurs, and 52 age– and sex-matched healthy controls. Participants were recruited from the general population via online and community advertisements. The study was approved by the University of Sydney Human Research Ethics Committees. All participants provided informed written consent in accordance with the Declaration of Helsinki.

Migraine diagnosis was established according to the criteria of the International Classification of Headache Disorders, 3^rd^ edition (ICHD-3) by neurologist R.J.S. Chronic migraine was defined as having a headache on 15 or more days per month for more than 3 months, and at least 8 days per month had features of migraine headache. Episodic migraine was defined as having a headache on less than 15 days per month for more than 3 months. Aura was defined as transient episodes of visual or sensory symptoms that develop gradually and are typically followed by a headache. Exclusion criteria for controls included any current pain symptoms, chronic pain conditions, use of analgesic medications, or history of neurological or psychiatric disorders. Migraineurs were excluded if they reported any comorbid neurological conditions or pain syndromes other than migraine. Among the 38 migraineurs, 20 participants (including 13 chronic migraineurs) reported migraine with aura.

### MRI acquisition

During MRI acquisition, 13 (72.2%) of the chronic migraineurs and 5 (25%) of the episodic migraineurs reported ongoing pain (Supplementary Table 1). High-resolution T1-weighted (T1w) MRI images were acquired using a 7 Tesla Plus Magnetom Siemens scanner (Magnetom Terra) equipped with a single channel transmit and 32-channel receive head coil (Nova Medical). A magnetisation-prepared rapid gradient-echo (MP2RAGE) sequence (Marques et al., 2010) was acquired with the following parameters: repetition time (TR) = 5000 ms, echo time (TE) = 2.04 ms, FA = 4°/5°, inversion time (TI1/T12) = 700/2700 ms, FOV=240 mm, matrix size=320×320, and voxel size = 0.75 mm isotropic.

### MRI preprocessing

PVS were automatically segmented from T1-weighted MRI scans using a previously validated deep learning model based on the nnU-Net residual encoder architecture (Isensee et al., 2021; Pham et al., 2025). The resulting PVS segmentation masks included unique voxel labels for PVS in the white matter and basal ganglia. The segmentations were used to compute total PVS volumes and cluster counts across the white matter and basal ganglia for each participant.

Brain volume extraction and quantification was performed using FastSurfer (v2) (Henschel et al., 2020). To assign PVS to white matter lobes and arterial vascular territories, segmentation masks were overlaid with two separate brain atlases. The brain extracted masks were spatially registered to the MNI152 standard space using the Advanced Normalization Tools (ANTs, v2.4.3) (Avants et al., 2011). Rigid, affine, and symmetric diffeomorphic normalisation were performed with antsRegistrationSyn.sh to create forward and inverse transforms. The transformations enabled the projection of PVS clusters onto two additional atlases. The ICBM2009c asymmetric white matter lobe atlas parcellates the frontal, parietal, temporal and occipital lobes (Fonov et al., 2009). An arterial vascular territory atlas defined the anterior cerebral artery (ACA), middle cerebral artery (MCA), and posterior cerebral artery (PCA) territories (Liu et al., 2023) (Figure 1). PVS were also quantified in the midbrain (MB) and hippocampus (HP) using region-specific nnU-Net models developed in prior work.

**Figure 1.**
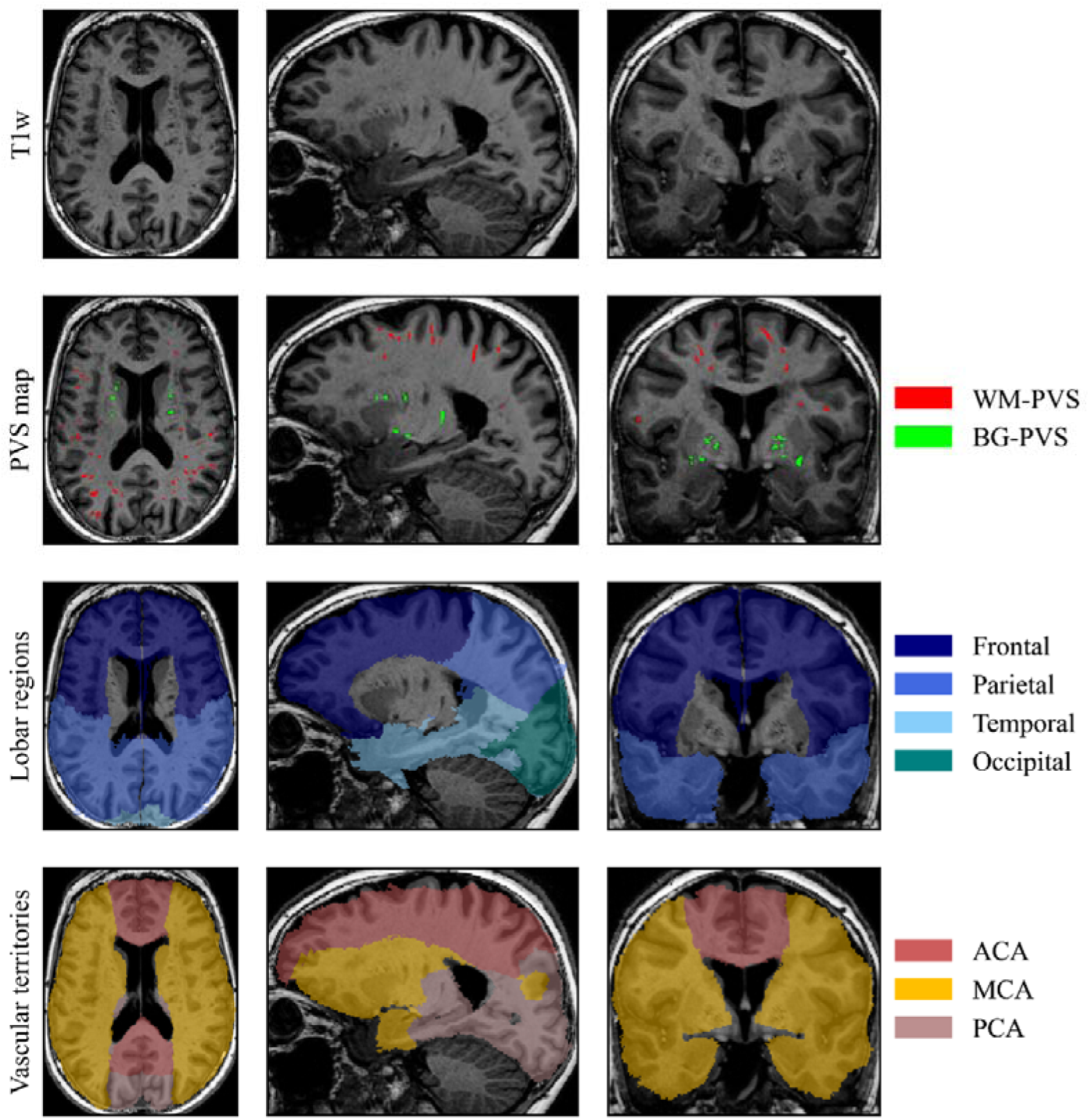
T1-weighted (T1w) MRI with segmented perivascular spaces (PVS) maps and region atlas parcellations according to lobar regions and arterial vascular territories. Top row: Raw T1w MRI scan from a patient with chronic migraine. Images from the axial, sagittal, and coronal planes are presented from left to right, respectively. Second row: PVS map overlaid on T1w MRI, with white matter PVS (WM-PVS, red) and basal ganglia PVS (BG-PVS, green). Third row: Lobar regions including the frontal (dark blue), parietal (light blue), temporal (cyan), and occipital (teal) lobes. Bottom row: Arterial vascular territories including the regions supplied by the anterior cerebral artery (ACA, brown), middle cerebral artery (MCA, yellow), and posterior cerebral artery (PCA, pink).

PVS clusters were assigned to each region of interest (ROI) based on the location of their centroids. Clusters centroids were calculated using the ‘measure.centroid’ function from the *scikit-image* Python library (v0.19.3; Python v3.9). A cluster was considered part of a region if its centroid fell within the ROI mask.

### Statistical analysis

Shapiro-Wilk tests were applied to assess the distribution of PVS metrics. As all PVS metrics significantly deviated from normality (*p*<0.05), group comparisons were analysed using generalised linear models (GLMs) with a Tweedie distribution and log-link function, deemed suitable for skewed data distributions. Separate GLMs were fitted for each PVS metric (volume and cluster count) within each ROI. All models included age, sex, and total brain volume (TBV) as covariates to account for inter-individual differences. These variables were selected based on prior work demonstrating their influence on PVS burden (Francis et al., 2019).

To evaluate the effect of migraine diagnosis, GLMs were constructed with group status (control vs migraine) as the main predictor. Additional models were fitted to assess differences between migraine subtypes (control vs episodic migraine, and control vs chronic migraine) and to test the effect of aura (presence vs absence) in migraineurs. *P*-values from all models were adjusted for multiple comparisons using the Benjamini-Hochberg false discovery rate (FDR) correction.

To validate automated PVS quantification, we compared the automated PVS counts with manual counts across multiple brain regions. In the centrum semiovale (CS) and basal ganglia (BG), comparisons were formed on two representative axial slices. Whereas in the midbrain and hippocampus, counts were assessed across the entire regions. Manual ratings were performed by a single experienced rater (WP) and independently reviewed by a radiologist (AJ). The evaluation was conducted on all MRI scans included in the study cohort. Agreement between automated and manual counts was assessed using Spearman’s rank correlation and Lin’s concordance correlation coefficients (CCC). Validation was performed for each region, including the centrum semiovale, basal ganglia, midbrain, and hippocampus. Statistical significance was defined as *p*<0.05. All statistical analyses were performed in R (v4.2.1) using the *EpiR* package for correlation and concordance analyses and the *glmmTMB* package for generalised linear models.

## Results

The study cohort included 90 participants (58 females, median [Q1, Q3] age=28.6 [25.1, 39.4] years), including 52 healthy controls (26 females, median age=26.9 [24.8, 37.6] years), 20 episodic migraineurs (17 females, age=28.6 [27.5, 32.3] years), and 18 chronic migraineurs (15 females, age=37.3 [27.9, 54] years) (Table 1). At the time of MRI acquisition, 13 chronic migraineurs and 5 episodic migraineurs were experiencing ongoing pain (Supplementary Table 1).

**Table 1.** Demographic and clinical characteristics of the study cohort. Numerical variables are presented as median [IQR]. PVS = perivascular space; TBV = total brain volume. Categorical variables were compared using the Chi-squared test, and continuous variables using non-parametric Kruskal-Wallis tests.

|  | Control (*n*=52) | Episodic Migraine (*n*=20) | Chronic Migraine (*n*=18) | Total (*N*=90) | *p*-value |
| --- | --- | --- | --- | --- | --- |
| **Age, years** | 26.9 [24.8, 37.6] | 28.6 [27.5, 32.3] | 37.3 [27.9, 54] | 28.6 [25.1, 39.4] | 0.08 |
| **Sex, *n* Female (%)** | 26 (50%) | 17 (85%) | 15 (83.3%) | 58 (64.4%) | 0.004 |
| **TBV (cm<sup>3</sup>)** | 1,100.7 [1,024, 1,141.1] | 1,058.7 [1,002.3, 1,098.4] | 1,056.2 [972.1, 1,104.4] | 1,078.2 [1,007.1, 1,124] | 0.11 |
| **Aura, *n* (%)** | – | 7 (35.0%) | 13 (72.2%) | 20 (52.6%) |  |
| **PVS volume (mm<sup>3</sup>)** | 1,486.1 [988.9, 2,526.1] | 1,339.9 [1,035.1, 1,878] | 1,268.2 [782.3, 2,342] | 1,445.6 [1,005.9, 2,322.7] | 0.58 |
| **PVS cluster counts** | 224.5 [180, 313] | 218.5 [169, 285.3] | 189 [139.5, 311.8] | 220.5 [166.3, 311.8] | 0.69 |

### Validation of automated perivascular space quantification

Perivascular space cluster counts in representative axial slices were compared between automated predictions and manual counts for the CS and BG. In the CS, automated counts showed strong correlation and moderate agreement with manual ratings (Spearman’s ρ=0.88, *p*<0.001; CCC=0.63, 95%CI [0.57, 0.68]). In contrast, no significant correlation and only negligible agreement were observed in the BG (Spearman’s ρ=0.02, *p*=0.75; CCC=0.05, 95%CI [0, 0.1]). For the midbrain and hippocampus, PVS counts demonstrated moderate consistency between methods. Correlation and agreement were modest in the midbrain (Spearman’s ρ=0.44, *p*<0.001; CCC=0.42, 95%CI [0.25, 0.57]) and hippocampal (Spearman’s ρ=0.55, *p*<0.001; CCC=0.51, 95%CI [0.36, 0.63]) regions. Consistency between automated and manual counts was comparable across regions in both controls and migraineurs (Table 2). Figure 2 presents examples of perivascular spaces identified in the white matter, basal ganglia, midbrain and hippocampus.

**Figure 2.**
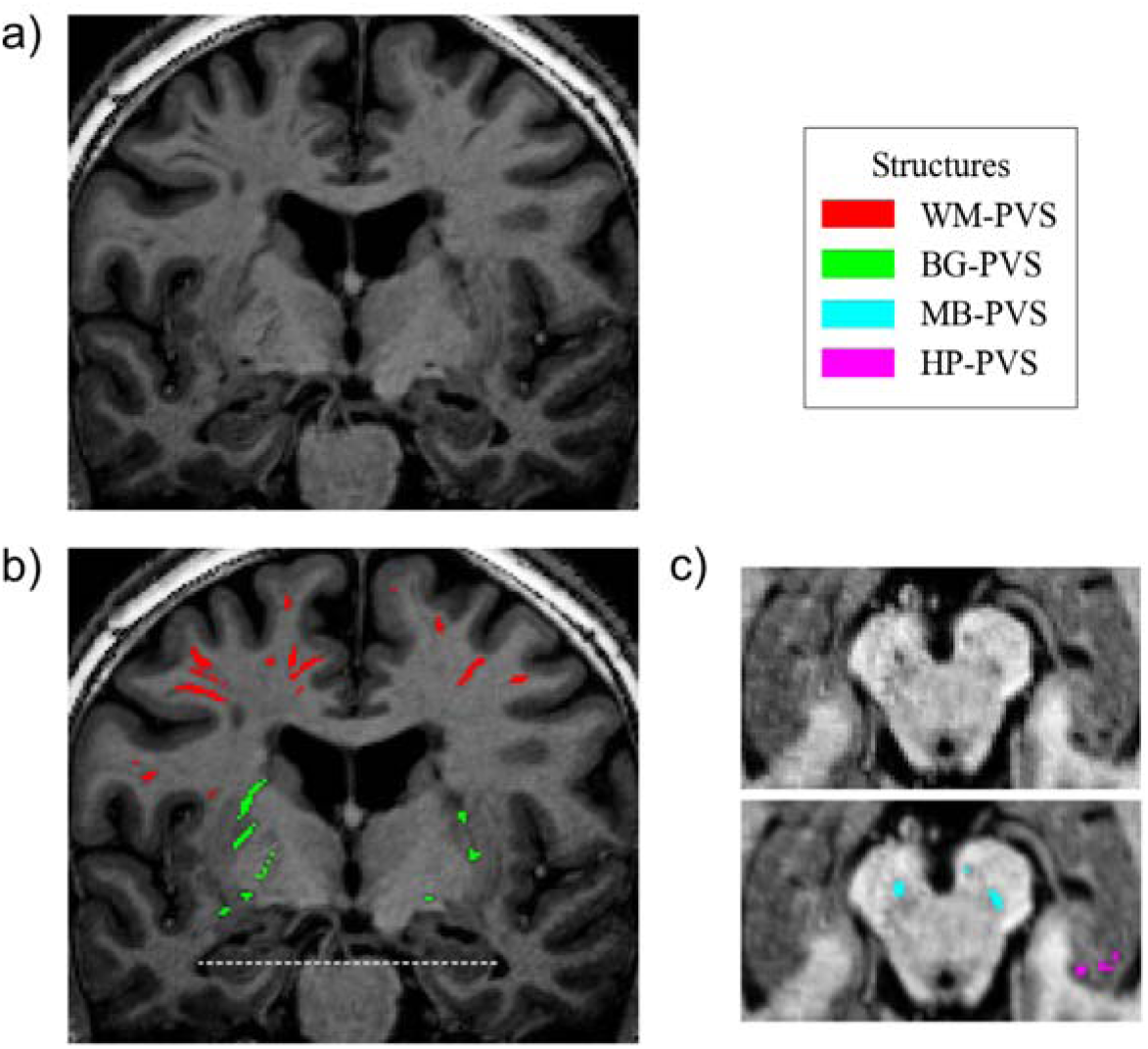
Perivascular spaces (PVS) identified in the white matter (WM, red), basal ganglia (BG, green), midbrain (MB, blue) and hippocampus (HP, purple) on a 7T T1-weighted MRI scan of an individual with chronic migraine. a) Coronal view of the raw T1-weighted image. b) Corresponding coronal slice with PVS segmentations overlaid. The white dashed line indicates the axial plane used for panel c. c) Axial view demonstrating PVS in the midbrain and hippocampus.

**Table 2.** Validation of automated perivascular space measurements across brain regions. Spearman’s correlation coefficients (ρ), Lin’s concordance correlation coefficients (CCC), and 95% CI between automated and manual PVS counts across regions of interest (ROI) for the entire cohort, controls, and migraineurs.

| Group | ROI | $\rho$ | *p*-value | CCC | 95%CI |
| --- | --- | --- | --- | --- | --- |
| **All** | CS | 0.88 | <0.001 | 0.63 | 0.57–0.68 |
|  | BG | 0.02 | 0.75 | 0.05 | 0–0.1 |
|  | MB | 0.44 | <0.001 | 0.42 | 0.25–0.57 |
|  | HP | 0.55 | <0.001 | 0.51 | 0.36–0.63 |
| **Controls** | CS | 0.91 | <0.001 | 0.65 | 0.58–0.71 |
|  | BG | -0.03 | 0.77 | 0.01 | -0.04–0.07 |
|  | MB | 0.36 | 0.001 | 0.39 | 0.15–0.58 |
|  | HP | 0.5 | <0.001 | 0.37 | 0.16–0.56 |
| **Migraine** | CS | 0.87 | <0.001 | 0.58 | 0.48–0.66 |
|  | BG | 0.05 | 0.70 | 0.1 | 0.01–0.18 |
|  | MB | 0.23 | 0.17 | 0.46 | 0.18–0.67 |
|  | HP | 0.62 | <0.001 | 0.74 | 0.57–0.85 |

### Impacts of migraine diagnosis, subtypes, and aura on perivascular spaces

When comparing all migraineurs to controls, migraineurs exhibited smaller PVS volumes in the midbrain (exp(β)=0.29, 95%CI [0.08, 0.73], *p*=0.02) and fewer PVS volumes in the basal ganglia (exp(β)=0.82, 95%CI [0.69, 1], *p*=0.04). No other regions differed significantly between controls and migraineurs.

In subgroup analyses, episodic migraineurs showed approximately 24% lower BG PVS volumes (exp(β)=0.76, 95%CI [0.61, 0.95], *p*=0.01) compared to controls. Episodic migraineurs demonstrated no additional regional differences in PVS measurements. In contrast, chronic migraineurs exhibited a broader pattern of PVS reduction relative to controls, including an approximately 18% decrease in whole-brain PVS cluster counts (exp(β)=0.82, 95%CI [0.68, 0.98], *p*=0.04). Regionally, PVS measures were 18–31% lower across the parietal (volume: exp(β)=0.73, 95%CI [0.56, 0.95], *p*=0.02; counts: exp(β)=0.8, 95%CI [0.68, 0.94], *p*=0.01) and temporal (volume: exp(β)=0.69, 95%CI [0.45, 1.02], *p*=0.06; counts: exp(β)=0.72, 95%CI [0.53, 0.96], *p*=0.03) lobes, and MCA territory (volume: exp(β)=0.78, 95%CI [0.61, 0.99], *p*=0.048; counts: exp(β)=0.82, 95%CI [0.68, 0.97], *p*=0.03) (Figure 3-4, Table 3). Chronic migraineurs also showed 82% reduction in midbrain PVS volumes compared to controls (exp(β)=0.18, 95%CI [0.02, 0.63], *p*=0.01).

**Figure 3.**
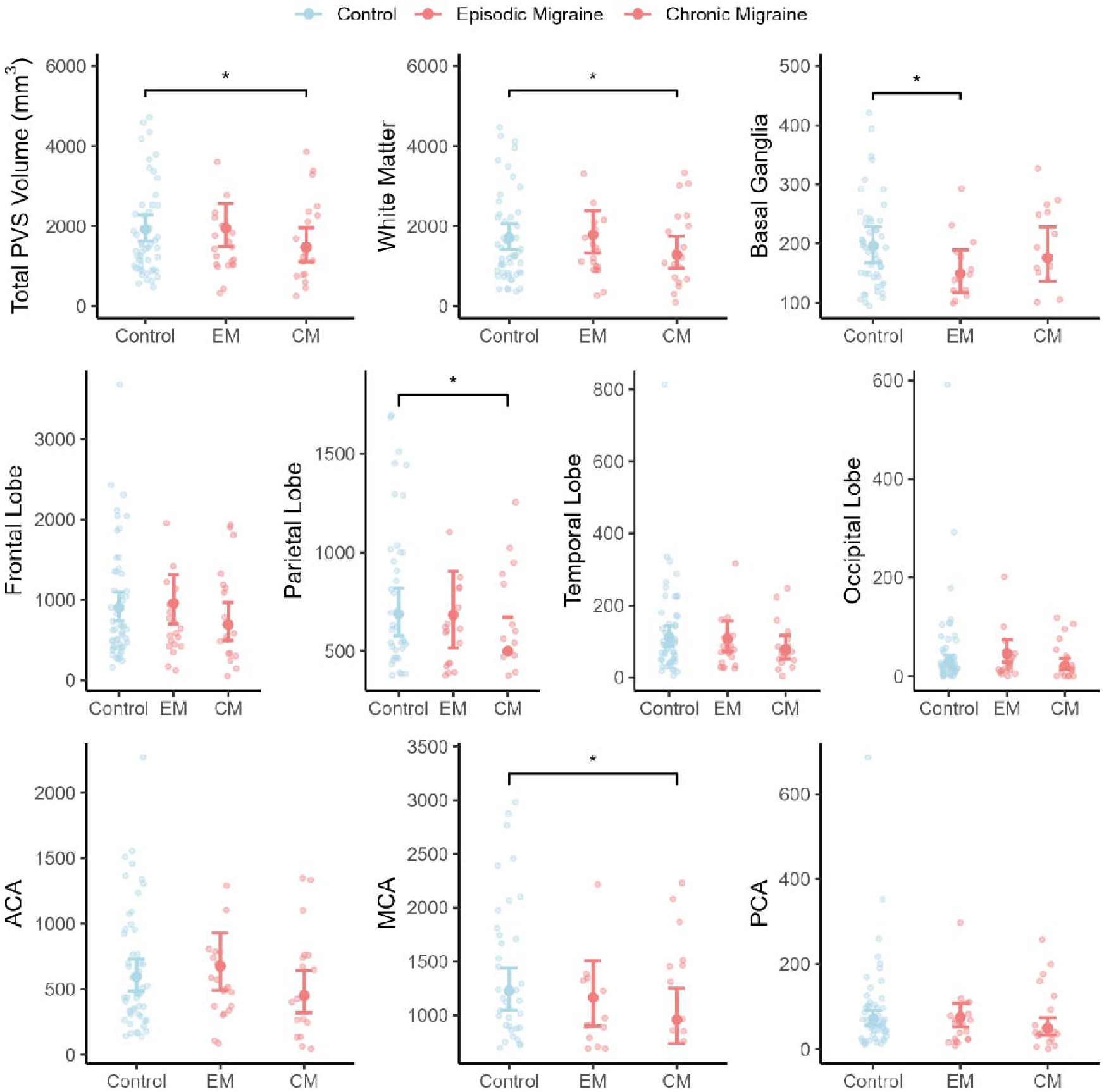
Group comparisons of perivascular space (PVS) volumes across brain regions. Estimated marginal means of PVS volumes (±95% CI) are shown for controls, episodic migraine (EM), and chronic migraine (CM) across the whole brain, white matter, basal ganglia, lobar regions (frontal, parietal, temporal, occipital), and vascular territories (anterior cerebral artery [ACA], middle cerebral artery [MCA], and posterior cerebral artery [PCA]). Notably, after corrections for multiple comparisons, none of these associations remained statistically significant (*p*_FDR_>0.05). Significance: \**p*<0.05.

**Figure 4.**
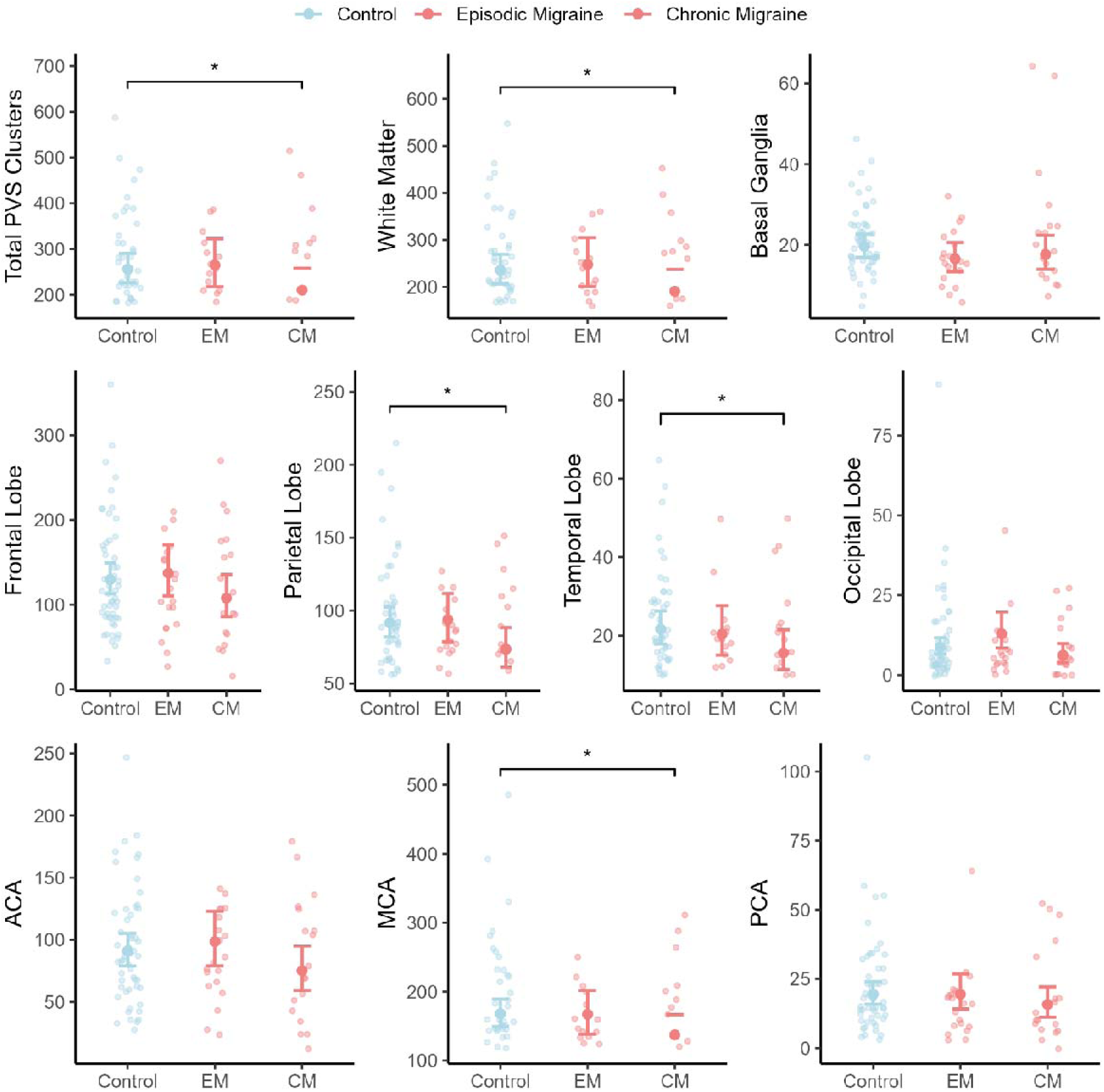
Group comparisons of perivascular space (PVS) cluster counts across brain regions. Estimated marginal means of PVS volumes (±95% CI) are shown for controls, episodic migraine (EM), and chronic migraine (CM) across whole brain, white matter, basal ganglia, lobar regions (frontal, parietal, temporal, occipital), and vascular territories (anterior cerebral artery [ACA], middle cerebral artery [MCA], and posterior cerebral artery [PCA]). Notably, after corrections for multiple comparisons, none of these associations remained statistically significant (*p*_FDR_>0.05). Significance: \**p*<0.05.

**Table 3.**
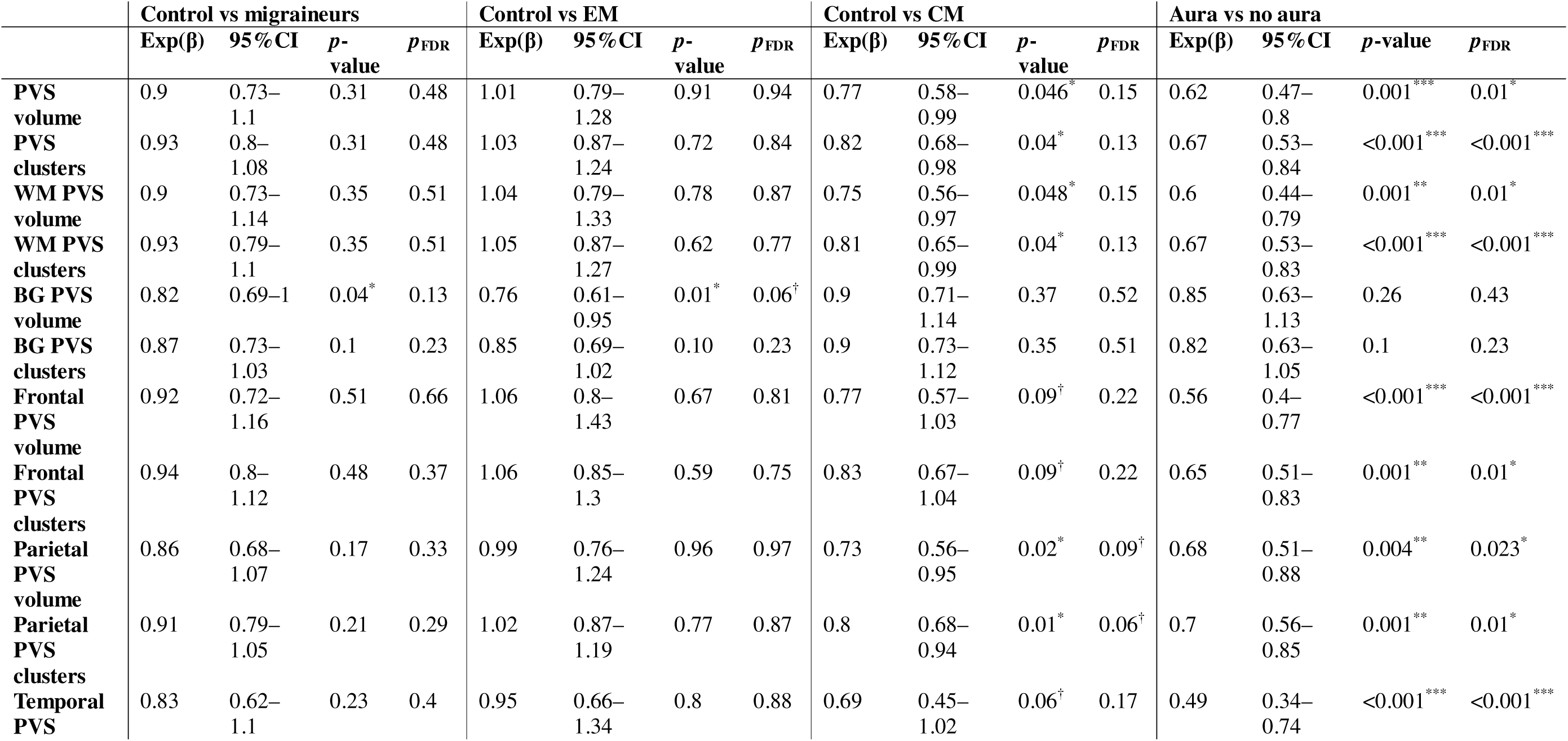
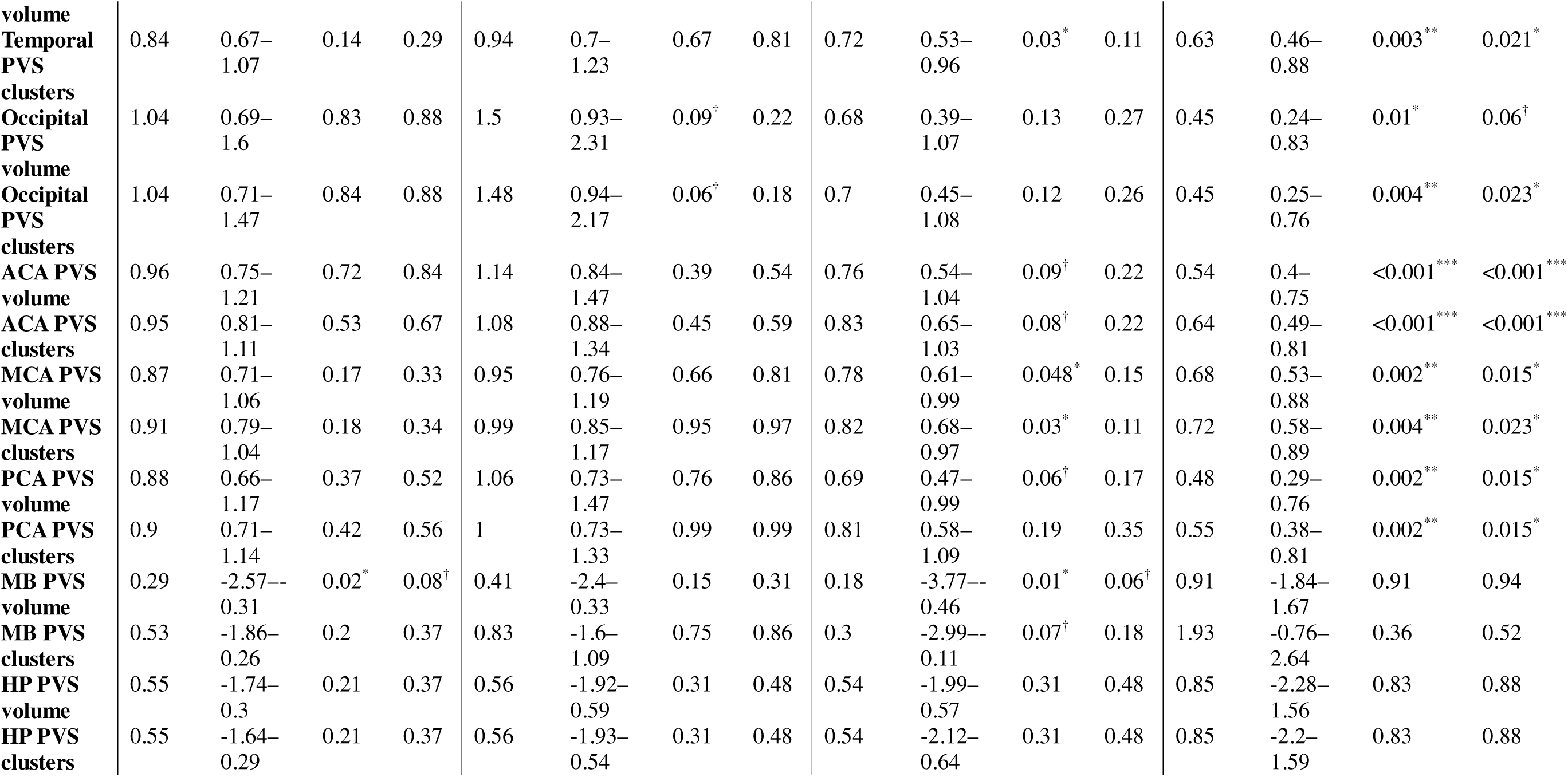
Results from generalised linear models assessing the effects of migraine diagnosis, migraine subtypes, and the presence of aura on perivascular space (PVS) measurements. Exponentiated β estimates, 95% CI’s, *p*-values, and FDR-corrected *p*-values are reported for comparisons of controls vs all migraineurs, controls vs episodic migraine (EM), controls vs chronic migraine (CM), and migraine with aura vs without aura. Outcomes include total PVS volume and cluster counts, as well as regional (white matter [WM], basal ganglia [BG], frontal, parietal, temporal, occipital lobes, and the anterior cerebral artery [ACA], middle cerebral artery [MCA], and posterior cerebral artery [PCA] territories). MB = midbrain; HP = hippocampus. All models were adjusted for age, sex, and total brain volume. Significance: ^†^p < 0.1, *p<0.05, **p<0.01, ***p<0.001.

After adjusting for confounders, migraine with aura was associated with widespread reductions in PVS measurements, ranging from approximately 28% to 55%, across the whole brain, white matter, all lobar regions, and major vascular territories (all *p*<0.05) (Figure 5-6, Table 3). In contrast, no associations were observed between the presence of aura and PVS in the basal ganglia, midbrain, or hippocampus.

**Figure 5.**
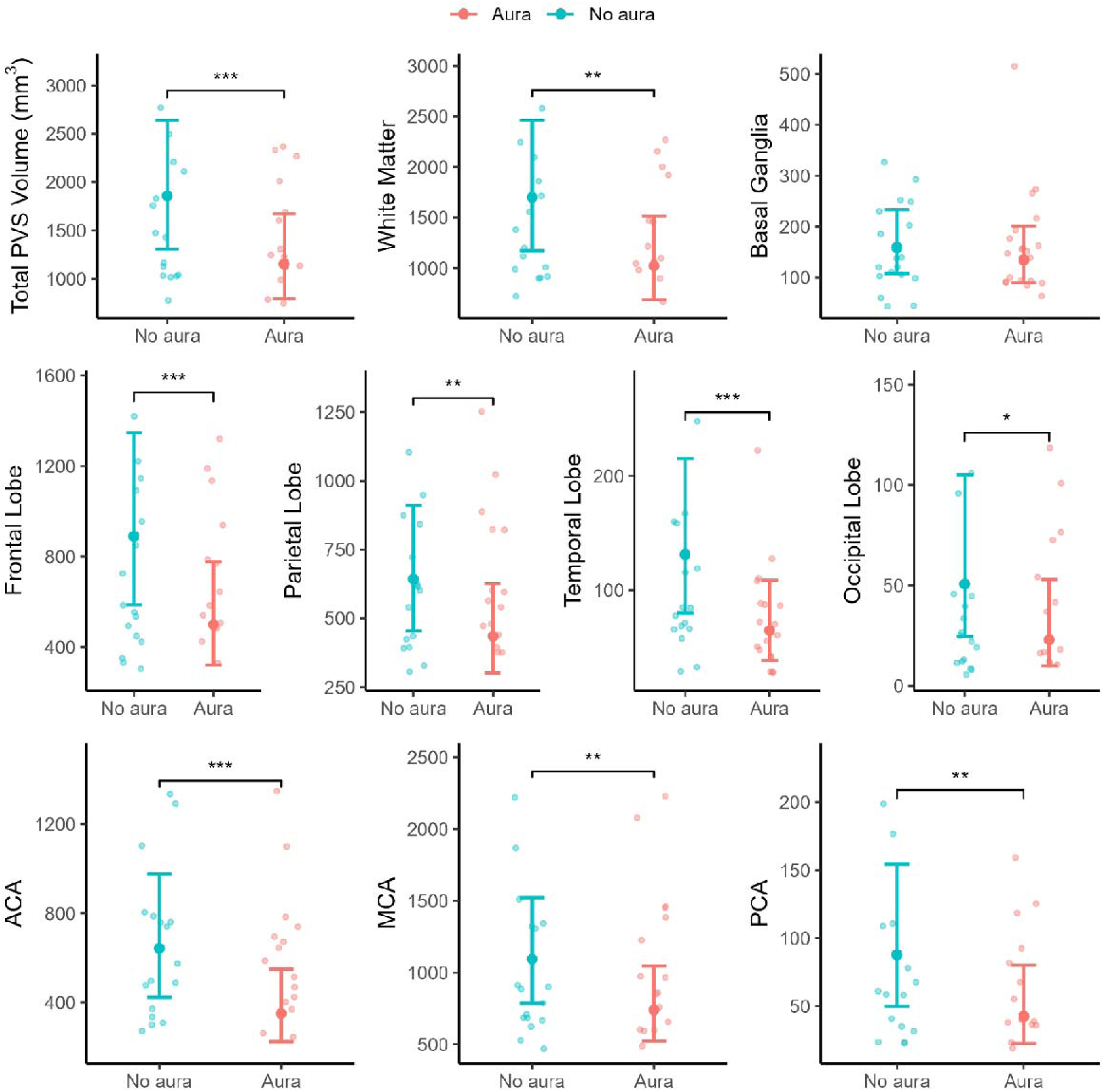
Comparisons of perivascular space (PVS) volumes between migraineurs with and without aura. Estimated marginal means (±95% CI) are displayed for PVS volumes across the whole brain, white matter, basal ganglia, lobar regions (frontal, parietal, temporal, occipital), and vascular territories (anterior cerebral artery [ACA], middle cerebral artery [MCA], and posterior cerebral artery [PCA]). All associations, except for PVS volumes in the occipital lobe, remained statistically significant after correction for multiple comparison (*p*_FDR_<0.05). Significance: \**p*<0.05, \*\**p*<0.01, \*\*\**p*<0.001.

**Figure 6.**
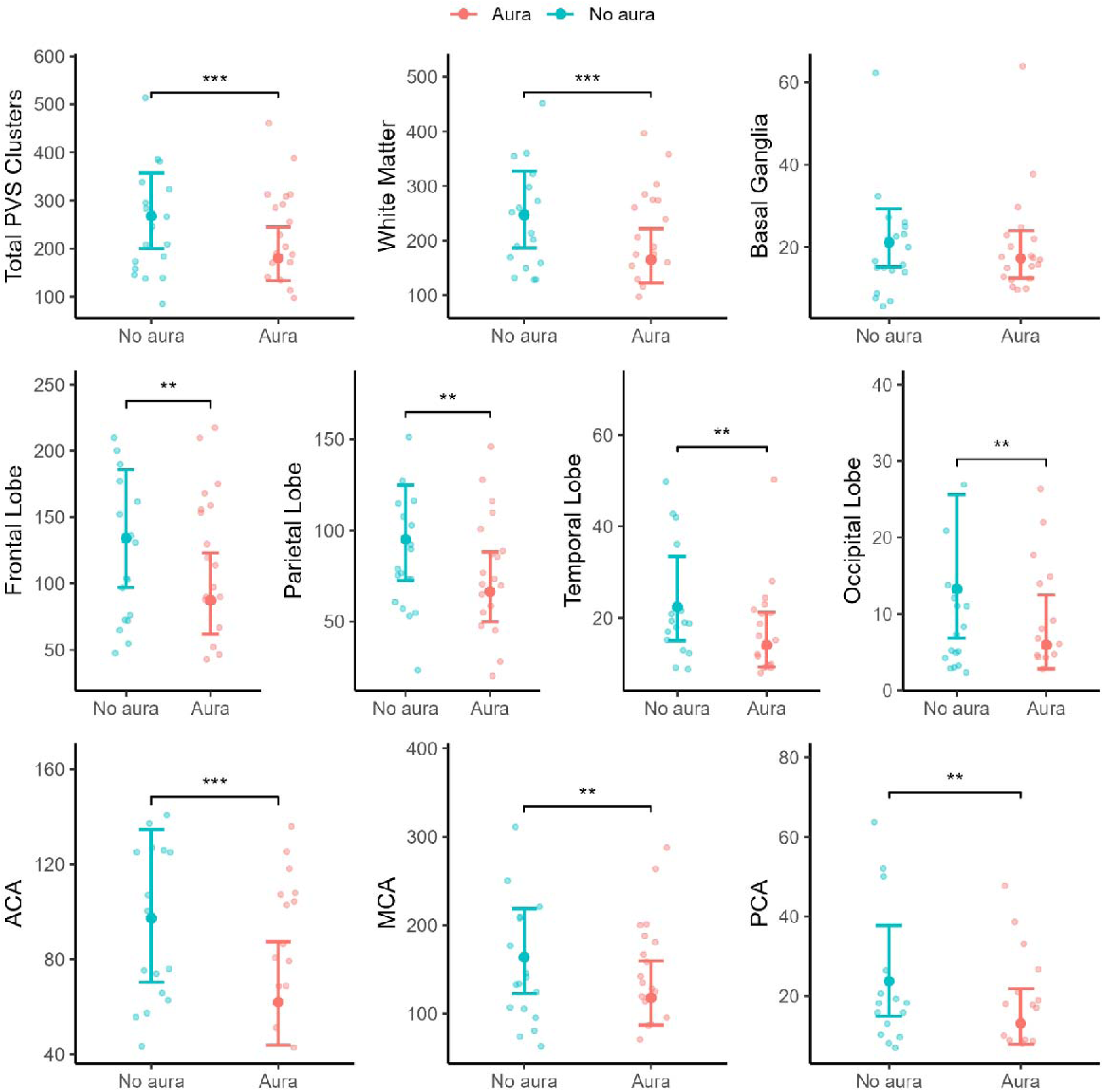
Comparisons of perivascular space (PVS) cluster counts between migraineurs with and without aura. Estimated marginal means (±95% CI) are displayed for PVS volumes across the whole brain, white matter, basal ganglia, lobar regions (frontal, parietal, temporal, occipital), and vascular territories (anterior cerebral artery [ACA], middle cerebral artery [MCA], and posterior cerebral artery [PCA]). All associations remained statistically significant after correction for multiple comparison (*p*_FDR_<0.05). Significance: \**p*<0.05, \*\**p*<0.01, \*\*\**p*<0.001.

After correction for multiple comparisons, only the associations between the presence of aura and PVS metrics across the whole brain, white matter, frontal, parietal, and temporal lobes, and major vascular territories remained statistically significant (*p*_FDR_<0.05). Associations between PVS metrics and either migraine diagnosis or migraine subtypes were no longer significant following adjustment.

## Discussion

This study identified distinct regional alterations in MRI-visible perivascular spaces in both episodic and chronic migraine. We observed distinct patterns of perivascular space alterations according to migraine diagnosis, suggesting that migraine frequency and chronification exert differential effects on brain fluid homeostasis and glymphatic function. In episodic migraine, reduced PVS volume was observed in the basal ganglia. By contrast, chronic migraine was characterised by reductions in PVS measurements across the parietal and temporal lobes, as well as in regions perfused by the middle cerebral artery. The presence of aura was linked to the most widespread alterations, with reduced PVS burden across nearly all brain regions except for the basal ganglia, midbrain, and hippocampus. These findings, enabled by high field 7T MRI and a state-of-the-art framework for automated PVS quantification, indicate that migraine chronicity and aura may significantly alter the perivascular pathways through which glymphatic flow occurs.

Contrary to previous MRI studies that reported PVS enlargement in migraine and headache populations, we observed a consistent pattern of PVS reduction. Earlier studies, often relying on visual rating scales or lower-resolution imaging, suggested increased PVS burden occurs in migraineurs, particularly in the centrum semiovale and midbrain (Yuan et al., 2023). In contrast, our findings align with evidence from animal models, where cortical spreading depression (the electrophysiological basis of migraine aura) has been shown to induce collapse of perivascular spaces and impair glymphatic inflow (Schain et al., 2017). Notably, previous animal studies using in vivo two-photon microscopy imaging have been constrained by shallowed imaging depth, limiting observations to superficial cortical regions. Our findings suggest that similar glymphatic disruptions may extend deeper into the white matter parenchyma in human migraine.

The physiological mechanisms underlying the observed reductions in PVS metrics remain unclear. One possibility is that cortical spreading depolarisation (CSD), which underlies migraine aura, induces sustained alterations in perivascular fluid dynamics. In rodent models, CSF triggers drastic but transient changes in perivascular fluid flow and morphology that typically resolve within an hour (Schain et al., 2017). In humans, however, CSF may produce more persistent structural and functional disturbances, making them MRI-detectable for a prolonged period of time. An alternative explanation is that secondary pathophysiological processes initiated by CSD, rather than the aura itself, drive these widespread reductions. For example, the release and distribution of vasoactive and neuroactive solutes triggered by CSD (Huang et al., 2023) may circulate across brain regions, disrupt perivascular fluid regulation, and impose lasting effects on PVS morphology and glymphatic function well beyond the initial aura event.

In episodic migraine, we did not observe significant PVS alterations in the defined vascular territories or lobar regions, apart from the occipital lobe. This may reflect a more complete recovery of glymphatic function between infrequent migraine attacks. However, we identified a significant reduction in basal ganglia PVS burden in episodic migraineurs. Given that BG-PVS enlargement is typically associated with hypertensive arteriopathy, the mechanism underlying reduced PVS in this region is unclear (Francis et al., 2019). One possibility is that CSF-induced disturbances initiate in cortical regions and gradually propagate into the deep subcortical grey matter, leading to delayed PVS alterations in deep brain structures. However, this remains speculative and should be addressed in future studies. Importantly, the lack of a significant correlation between manual PVS counts in representative axial slices of the basal ganglia and the corresponding automated BG-PVS counts suggests that the observed association between reduced BG-PVS in episodic migraine may reflect a spurious finding.

The discrepancies between our findings and prior PVS studies in migraine are likely attributable to methodological differences. Our study differs in several key aspects: the use of high field 7T MRI, automated segmentation of PVS, and regional analyses. Previous studies used lower field strength MRI, subjective rating scales, and investigated paediatric or general headache populations (Husøy et al., 2016; Jeon et al., 2023; Yuan et al., 2023).

## Limitations

This study has several limitations. The modest sample size may have limited statistical power to detect additional differences in PVS burden, particularly in the regions such as the midbrain, where PVS alterations have previously been reported in a migraine cohort. The cross-sectional design allowed for the identification of between-group differences but precludes any inference about the temporal dynamics of PVS alterations. It remains unclear whether the observed differences represent transient features associated with migraine episodes or sustained structural changes. Longitudinal imaging is needed to address this question.

Sleep is a major regulator of brain waste clearance and migraine pathophysiology (Chong et al., 2022; Vgontzas & Pavlović, 2018). Although sleep was not assessed in the present study, migraine symptoms and brain-wide physiological changes are intertwined with sleep disturbances. The lack of sleep-related measurements therefore limits our ability to interpret the functional significance of the observed PVS alterations, as some of these changes may be partially driven by disrupted sleep. Notably, the strongest associations in our study linked the presence of aura, a phenomenon for which there is relatively little evidence of major sleep-related effects. This raises the possibility that aura-related PVS changes may reflect mechanisms beyond disruptions in sleep. Given the importance of sleep and its prevalence in migraine disorders, future studies should examine the interplay between sleep characteristics, PVS morphology on MRI, and aura (Vgontzas & Pavlović, 2018). The integration of multimodal assessments, including sleep monitoring and CSF flow measurements, would provide useful for understanding the relevance of perivascular spaces in migraine pathophysiology.

Although our use of high field 7T MRI enabled precise delineation of PVS morphology, the clinical generalisability of these findings is limited. At present, 7T is primarily a research tool, and it remains uncertain whether similar results could be replicated using standard 3T or 1.5T scanners. Given the novelty of our findings and their divergence from prior reports, replication in larger cohorts and across multiple MRI field strengths is essential to confirm their veracity.

Additionally, our imaging protocol did not include fluid-attenuated inversion recovery (FLAIR) sequences, which limited our ability to differentiate PVS from confluent white matter hyperintensities (WMHs). This is particularly relevant given the known prevalence of WMHs in migraineurs (Hamedani, 2013; Kruit et al., 2010; Zhang et al., 2024). The presence of WMHs may have obscured PVS, potentially influencing the accuracy of our findings. Although this study focused on T1w MRI, future research incorporating T2-weighted MRI, which provides superior contrast for visualising PVS, may enable more accurate delineation and quantification (Duering et al., 2023; Wardlaw et al., 2013).

## Conclusion

This study provides novel evidence that migraine, particularly in its chronic form, is associated with region-specific reductions in MRI-visible perivascular spaces. Chronic migraineurs exhibited diminished PVS volume and cluster counts across widespread white matter regions and vascular territories, suggesting cumulative disruption to brain-wide fluid transport. Whereas episodic migraine was selectively associated with reduced PVS in the basal ganglia, highlighting potential regional vulnerability with less frequent migraine episodes. These findings are consistent with the hypothesis that migraine involves impaired glymphatic function, concurrent with perivascular space collapse associated with migraine aura. MRI-PVS may therefore serve as a non-invasive imaging marker for glymphatic dysfunction driven by migraine frequency. Future studies incorporating longitudinal designs, multi-modal imaging, or sleep metrics may further elucidate the interplay between migraine, glymphatic clearance, and alterations in perivascular spaces.

## Data Availability

The dataset analysed in the current study are available from the corresponding author upon reasonable request.

## Acknowledgements

We would like to thank the volunteers for their involvement in the study. We thank R. Glarin for her assistance with image protocol development and acquisition. The authors acknowledge the facilities and scientific and technical assistance from the National Imaging Facility, a National Collaborative Research Infrastructure Strategy capability, at the Melbourne Brain Centre Imaging Unit, The University of Melbourne. This work was supported by a research collaboration agreement with Siemens Healthineers.

## References

1. Avants, B. B., Tustison, N. J., Song, G., Cook, P. A., Klein, A., & Gee, J. C. (2011). A reproducible evaluation of ANTs similarity metric performance in brain image registration. NeuroImage, 54(3), 2033–2044. 10.1016/j.neuroimage.2010.09.025

2. Ayata, C., & Lauritzen, M. (2015). Spreading Depression, Spreading Depolarizations, and the Cerebral Vasculature. Physiological Reviews, 95(3), 953–993. 10.1152/physrev.00027.2014

3. Chong, P. L. H., Garic, D., Shen, M. D., Lundgaard, I., & Schwichtenberg, A. J. (2022). Sleep, cerebrospinal fluid, and the glymphatic system: A systematic review. Sleep Medicine Reviews, 61. 10.1016/j.smrv.2021.101572

4. Duering, M., Biessels, G. J., Brodtmann, A., Chen, C., Cordonnier, C., De Leeuw, F.-E., Debette, S., Frayne, R., Jouvent, E., Rost, N. S., Ter Telgte, A., Al-Shahi Salman, R., Backes, W. H., Bae, H.-J., Brown, R., Chabriat, H., De Luca, A., deCarli, C., Dewenter, A., … Wardlaw, J. M. (2023). Neuroimaging standards for research into small vessel disease—Advances since 2013. The Lancet Neurology, 22(7), 602–618. 10.1016/S1474-4422(23)00131-X

5. Ferrari, M. D., Goadsby, P. J., Burstein, R., Kurth, T., Ayata, C., Charles, A., Ashina, M., Van Den Maagdenberg, A. M. J. M., & Dodick, D. W. (2022). Migraine. Nature Reviews Disease Primers, 8(1), 2. 10.1038/s41572-021-00328-4

6. Fonov, V., Evans, A., McKinstry, R., Almli, C., & Collins, D. (2009). Unbiased nonlinear average age-appropriate brain templates from birth to adulthood. NeuroImage, 47, S102. 10.1016/s1053-8119(09)70884-5

7. Francis, F., Ballerini, L., & Wardlaw, J. M. (2019). Perivascular spaces and their associations with risk factors, clinical disorders and neuroimaging features: A systematic review and meta-analysis. International Journal of Stroke, 14(4), 359–371. 10.1177/1747493019830321

8. Hamedani, A. G. (2013). Migraine and white matter hyperintensities.

9. Henschel, L., Conjeti, S., Estrada, S., Diers, K., Fischl, B., & Reuter, M. (2020). FastSurfer—A fast and accurate deep learning based neuroimaging pipeline. NeuroImage, 219. 10.1016/j.neuroimage.2020.117012

10. Huang, W., Zhang, Y., Zhou, Y., Zong, J., Qiu, T., Hu, L., Pan, S., & Xiao, Z. (2023). Glymphatic Dysfunction in Migraine Mice Model. Neuroscience, 528, 64–74. 10.1016/j.neuroscience.2023.07.027

11. Husøy, A. K., Indergaard, M. K., Honningsvåg, L. M., Håberg, A. K., Hagen, K., Linde, M., Gårseth, M., & Stovner, L. J. (2016). Perivascular spaces and headache: A population-based imaging study (HUNT-MRI). Cephalalgia, 36(3), 232–239. 10.1177/0333102415587691

12. Isensee, F., Jaeger, P. F., Kohl, S. A. A., Petersen, J., & Maier-Hein, K. H. (2021). nnU-Net: A self-configuring method for deep learning-based biomedical image segmentation. Nature Methods, 18(2), 203–211. 10.1038/s41592-020-01008-z

13. Jeon, C. W., Lim, G. Y., & Moon, J. U. (2023). Dedicated neuroimaging analysis in children with primary headaches: Prevalence of lesions and a comparison between patients with and without migraines. 1–7.

14. Kruit, M., Van Buchem, M., Launer, L., Terwindt, G., & Ferrari, M. (2010). Migraine is associated with an increased risk of deep white matter lesions, subclinical posterior circulation infarcts and brain iron accumulation: The population-based MRI CAMERA study. Cephalalgia, 30(2), 129–136. 10.1111/j.1468-2982.2009.01904.x

15. Liu, C. F., Hsu, J., Xu, X., Kim, G., Sheppard, S. M., Meier, E. L., Miller, M. I., Hillis, A. E., & Faria, A. V. (2023). Digital 3D Brain MRI Arterial Territories Atlas. Scientific Data, 10(1), 1–17. 10.1038/s41597-022-01923-0

16. Marques, J. P., Kober, T., Krueger, G., van der Zwaag, W., Van de Moortele, P.-F., & Gruetter, R. (2010). MP2RAGE, a self bias-field corrected sequence for improved segmentation and T1-mapping at high field. NeuroImage, 49(2), 1271–1281. 10.1016/j.neuroimage.2009.10.002

17. Pham, W., Jarema, A., Rim, D., Chen, Z., Khlif, M. S. H., Macefield, V. G., Henderson, L. A., & Brodtmann, A. (2025). *A Comprehensive Framework for Automated Segmentation of Perivascular Spaces in Brain MRI with the nnU-Net* (arXiv:2411.19564). arXiv. 10.48550/arXiv.2411.19564

18. Pham, W., Lynch, M., Spitz, G., O’Brien, T., Vivash, L., Sinclair, B., & Law, M. (2022). A critical guide to the automated quantification of perivascular spaces in magnetic resonance imaging. Frontiers in Neuroscience, 16(December), 1–27. 10.3389/fnins.2022.1021311

19. Pietrobon, D., & Moskowitz, M. A. (2014). Chaos and commotion in the wake of cortical spreading depression and spreading depolarizations. Nature Reviews Neuroscience, 15(6), 379–393. 10.1038/nrn3770

20. Schain, A. J., Melo-Carrillo, A., Strassman, A. M., & Burstein, R. (2017). Cortical spreading depression closes paravascular space and impairs glymphatic flow: Implications for migraine headache. Journal of Neuroscience, 37(11), 2904–2915. 10.1523/JNEUROSCI.3390-16.2017

21. Schick, S., Gahleitner, A., Wöber-Bingöl, C., Wöber, C., Ba-Ssalamah, A., Schoder, M., Schindler, E., & Prayer, D. (1999). Virchow-Robin spaces in childhood migraine. Neuroradiology, 41(4), 283–287. 10.1007/s002340050749

22. Vgontzas, A., & Pavlović, J. M. (2018). Sleep Disorders and Migraine: Review of Literature and Potential Pathophysiology Mechanisms. Headache: The Journal of Head and Face Pain, 58(7), 1030–1039. 10.1111/head.13358

23. Wardlaw, J. M., Smith, E. E., Biessels, G. J., Cordonnier, C., Fazekas, F., Frayne, R., Lindley, R. I., O’brien, J. T., Barkhof, F., Benavente, O. R., Black, S. E., Brayne, C., Breteler, M., Chabriat, H., Decarli, C., De Leeuw, F.-E., Doubal, F., Duering, M., Fox, N. C., … Dichgans, M. (2013). Neuroimaging standards for research into small vessel disease and its contribution to ageing and neurodegeneration. Lancet Neurol, 12(8), 822–838. 10.1016/S1474-4422(13)70124-8

24. Yuan, Z., Li, W., Tang, H., Mei, Y., Qiu, D., Zhang, M., Sun, Q., Wang, W., Zhang, P., Ma, Z., Zhang, X., Zhang, Y., Wang, Y., & Yu, X. (2023). Enlarged perivascular spaces in patients with migraine: A case–control study based on 3T MRI. Annals of Clinical and Translational Neurology, 10(7), 1160–1169. 10.1002/acn3.51798

25. Zhang, Y., Li, Y., & He, L. (2024). Correlation between migraine and cerebral small vessel disease: A case–control study. European Journal of Pain, 28(4), 551–564. 10.1002/ejp.2199

