## Supplementary Table 1 for "MRI-Perivascular Spaces in Chronic and Episodic Migraine Disorder"

**Supplementary Table 1.** Chronic migraine (CM) and episodic migraine (EM) participant characteristics. Table includes age range, sex (F – female, M – male), head pain side (R – right, L – left, B – bilateral), aura (Y – yes, N – no), number of migraine and headache days per month, migraine pain intensity, regular medications, and medication and presence of pain during magnetic resonance imaging (MRI) scan.

|  |  |  |  |  |  |  |  |  | **During scan** | |
| --- | --- | --- | --- | --- | --- | --- | --- | --- | --- | --- |
| **Subject** | **Diagnosis** | **Age** | **Sex** | **Pain side** | **Aura** | **Migraine/headache days per month** | **Migraine pain intensity range** | **Regular medication** | **Medication** | **Pain/no pain** |
| 1 | CM | 26-30 | F | B | Y | 5-6 headaches,14-17 migraines | 2-4 | Contraceptive Pill Estelle (daily) | Contraceptive Pill Estelle | No Pain |
| 2 | CM | 51-55 | M | B | Y | 3-5 headaches/2-3 migraines | 1-3 | None | None | No Pain |
| 3 | CM | 21-25 | F | L | Y | Daily (8-12 headaches,16-20 migraines) | 5 | Propranolol 80mg (daily) | None | No Pain |
| 4 | CM | 31-35 | F | B | Y | Daily (27 headaches, 15 migraines) | 1-5 | Isotretinoin | None | Pain (head) |
| 5 | CM | 66-70 | F | R | Y | 11 headaches/migraines | 2-5 | Botox  Inderal 80mg daily  Perindopril 5mg | None | No Pain |
| 6 | CM | 36-40 | F | R | Y | 5 headaches/2 migraines | 3-5 | Pristiq 200mg daily | None | No Pain |
| 7 | CM | 56-60 | F | B | Y | 18 headaches, 12 migraines | 2-4 | Propranolol 80mg  Migraine care (daily) | Propranolol | Pain (head) |
| 8 | CM | 21-25 | F | R | N | 9 headaches, 8 migraines | 2-4 | Galcanezumab (monthly)  Magnesium (daily)  Lisdexamfetamine 60mg (daily)  Occasional: Fexofenadine and Doxylamine succinate | Lisdexamfetamine 50mg  Propranolol 20mg | Pain (neck) |
| 9 | CM | 26-30 | M | B | Y | 28 migraines | 2-5 | None | None | Pain (head) |
| 10 | CM | 51-55 | M | L | Y | 15 migraines | 1-5 | Duloxatine 120mg  Olmesartan 40mg | Fluoxetine 40mg  Olmesartan 40mg | Pain (head) |
| 11 | CM | 36-40 | F | R | Y | 10-15 headaches, 10-15 migraines | 3-5 | Amitriptyline  Levothyroxine | Levothyroxine 50mg | Pain (hip) |
| 12 | CM | 55-60 | F | B | Y | 27 headaches, 3-4 migraines | 2-4 | Triptan 50mg PRN, Asprin soluable 1g  PRN, Metoclopramide 10mg PRN, Propranolol (2x daily) | Propranolol | Pain (head) |
| 13 | CM | 26-30 | F | L | Y | Daily (constant headaches, 10-15 migraines) | 2-5 | Microgynon  Fexofenadine  Clonidine | Levest Fexofenadine  Clonidine | Pain (head) |
| 14 | CM | 21-25 | F | B | Y | Daily migraine | 3-5 | Contraceptive Pill | None | Pain (head) |
| 15 | CM | 31-35 | F | variable | N | Daily (23 headaches, 8 migraines) | 4-5 | Pizotifen 500mcg (daily)  Contraceptive Pill  Ambien  Melatonin  Clonzapam | None | Pain (head) |
| 16 | CM | 46-50 | F | unilateral (did not specify side) | N | Daily (23 headaches, 23 migraines) | 4-5 | Antidepressants | None | Pain (head) |
| 17 | CM | 36-40 | F | variable | Y | Daily (25-30 headaches, 20 migraines) | 2-5 | Botox  Levothyroxine Antidepressant | Escitalopram | Pain (head) |
| 18 | CM | 51-55 | F | L | N | Daily (constant headaches daily, 20 migraines) | 1-4 | CBD Oil 0.5ml (twice daily)  HRT  Gabapentin | Telmisartan  HRT  CBD Oil | Pain (head) |
| 19 | EM | 26-30 | F | L | N | 4 migraines, 3 headaches | 2-4 | None | None | No Pain |
| 20 | EM | 31-35 | F | R | N | 4 migraines | 1-3 | Carbimazole  Escitalopram Propranolol | None | No Pain |
| 21 | EM | 31-35 | M | R | N | 5 migraines, 1 headache | 1-3 | Albuterol | None | No Pain |
| 22 | EM | 36-40 | F | R | N | 4-7 migraines | 2-4 | None | None | Pain  (Migraine) |
| 23 | EM | 61-65 | F | L | Y | 4 migraines, 2 headaches | 2-4 | Amitriptyline 35mg  Pregabalin 25mg | Pregabalin 35mg  Amitriptyline | Pain (in feet) |
| 24 | EM | 26-30 | F | R | Y | Less than 1 migraine, 5 headaches | 3-5 | Fluoxetine | Fluoxetine 20mg | Pain (Headache, shoulder pain) |
| 25 | EM | 26-30 | M | L | N | 1 migraine, 2 headaches, | 2-3 | Temazepan as needed | None | No Pain |
| 26 | EM | 26-30 | F | R | N | 5 migraines, 6 headaches | 1-4 | None | None | No Pain |
| 27 | EM | 41-45 | F | R | N | 2 migraines, 3 headaches | 3-4 | None | None | No Pain |
| 28 | EM | 26-30 | F | B | Y | 6 migraines, 10 headaches | 2-4 | Preventative: Ajovy, lyrica, propranolol. Abortive: Sumatriptan, naproxen, ondansetron, maxalon, naramig | Ondansetron | Pain (head, neck, back) |
| 29 | EM | 26-30 | M | B | N | 5 migraines, 6 headaches | 3-5 | Nortriptyline 25mg  Sertraline 100mg | Sertraline 100mg | No Pain |
| 30 | EM | 26-30 | M | B | N | 3 migraines, 4 headaches | 3-4 | Escitalopram | None | No Pain |
| 31 | EM | 21-25 | F | L | N | 1 migraine, 3-4 headaches | 2-5 | None | None | No Pain |
| 32 | EM | 21-25 | F | L | N | 2 migraines, 15 headaches | 2-4 | Pizotifen 1500mcg  Methylphenidate  Cyclosporin  Melatonin  Clonidine | None | Pain (migraine) |
| 33 | EM | 26-30 | F | B | Y | 3 migraines, 7 headaches | 4-5 | None | None | No Pain |
| 34 | EM | 26-30 | F | L | N | 2 migraines | 2-5 | Propranolol 40mg daily  Isotretinoin | Propranolol  Antihistamine | No Pain |
| 35 | EM | 51-55 | F | L | Y | 1 migraine, 5 headaches | 1-5 | Vitamin B2 daily  Propranolol daily  Statin  Levothyroxine Calcium  Vitamin D | None | No Pain |
| 36 | EM | 31-35 | F | L | Y | 1 migraine, 4 headaches | 2-4 | Escitalopram 20mg | Aspirin | No Pain |
| 37 | EM | 31-35 | M | Not specified | Y | 2-3 migraines, 1 headache | 2-3 | None | None | No Pain |
| 38 | EM | 21-25 | F | B | N | 3 migraines, 2 headaches | 2-5 | None | Sulfasalazine 500mg x 2  Ursodeoxycholic acid 250mg x 3  Sertraline 50mg  Tioguanine 20mg  Cal-600 | No Pain |
